# Prevalence, Concordance, and Related Factors of Four Cardiovascular Risk Calculators in the Peruvian Population: Analysis of a National Survey

**DOI:** 10.1101/2024.05.22.24307712

**Authors:** Víctor Juan Vera-Ponce, Fiorella E. Zuzunaga-Montoya, Luisa Erika Milagros Vásquez-Romero, Joan A. Loayza-Castro, Enrique Vigil-Ventura, Carmen Inés Gutierrez De Carrillo

## Abstract

**Introduction:** Cardiovascular risk calculators (CVR) are essential for effectively preventing and managing cardiovascular diseases.

**Objective:** To determine the prevalence, the concordance among different CVR calculators, and the factors associated with these in the Peruvian population.

**Methods:** Analysis of data collected by the Food and Nutritional Surveillance Survey by Life Stages. The CVR calculators used were the AHA/ASCVD, World Health Organization (WHO) laboratory-based and non-laboratory-based, and Framingham. The percentage of risk for each individual in the study was calculated, categorizing the CVR of each calculator into low (<10%) vs moderate/high (≥10%). Concordance was analyzed through the kappa index.

**Results:** The level of moderate/high CVR risk according to the type of calculator used was 5.44%, 4.73%, 6.62%, and 23.88% for WHO laboratory-based, non-laboratory-based, AHA/ASCVD, and Framingham, respectively. Common related factors identified were male gender, age between 50–59 years, current smoker, hypertension, and diabetes. WHO laboratory-based and AHA/ASCVD showed higher concordance (kappa of 0.7289). In contrast, concordance between WHO laboratory-based and Framingham and between WHO non-laboratory-based and Framingham was relatively lower (kappa of 0.3098 and 0.2732, respectively).

**Conclusions:** There are discrepancies in CVR levels according to the different calculators evaluated, especially with Framingham, and common associated factors were identified. These variations suggest that prediction tools might not be universally applicable in their current form, making it essential to consider calibrating existing risk calculators or developing new tools that more accurately reflect the epidemiological and health profiles of the Peruvian population.

## Introduction

Cardiovascular disease (CVD) represents one of the leading causes of morbidity and mortality worldwide, posing a significant challenge to public health systems. The World Health Organization (WHO) estimates that cardiovascular diseases cause roughly 17.9 million deaths per year, accounting for approximately 31% of all fatalities worldwide. Therefore, preventing and effectively managing CVD through early identification of individuals at risk is crucial. In this context, cardiovascular risk calculators (CVR) are essential risk stratification and clinical decision-making tools. Their application in clinical practice facilitates informed decision-making regarding interventions such as lifestyle changes and pharmacological treatments ^(1,2)^.

While various studies have highlighted the utility of CVR calculators in predicting cardiovascular events, a fundamental question arises regarding their universal applicability across different populations ^(3–5)^. This question focuses on whether these tools, despite their varied methodologies and variables, offer consistent predictions for individuals of various ethnic backgrounds, ages, and socioeconomic conditions. The existence of multiple calculators, such as the AHA/ASCVD (American Heart Association/American College of Cardiology Atherosclerotic Cardiovascular Disease) ^(6)^, WHO laboratory-based and non-laboratory ^(7)^, and Framingham ^(8)^, with their respective sets of variables, suggests that they should provide similar risk estimates for a given patient. However, the reality is that a patient may be classified into different risk categories depending on the calculator used. This gap calls into question the precision and generalizability of such instruments. It highlights the importance of a meticulous cross-cultural assessment to decide their dependability and relevance when considering different groups ^(4,9–12)^.

Therefore, by implementing a national survey, the present study aims to determine the prevalence and concordance among different CVR calculators, and the factors related to them in the Peruvian population.

## Methods

### Study Design and Context

This is a cross-sectional and secondary analysis of data collected by the Life Stage Nutritional Surveillance Survey (VIANEV), implemented during 2017-2018 by Peru’s National Center for Food and Nutrition (CENAN) ^(13)^. This manuscript adheres to the STROBE (Strengthening the Reporting of Observational Studies in Epidemiology) guidelines to ensure the study’s robustness and transparency ^(14)^.

### Population, Sample, and Eligibility Criteria

The study utilized VIANEV data, which encompasses three distinct areas: the metropolitan area of Lima, other urban areas, and rural sectors throughout the country. The data collection procedure was based on a sampling scheme incorporating stratification, multiple stages, and a probabilistic approach, operating independently in each area. The sample selection was conducted in two main phases: first, by randomly choosing groupings of primary sampling units, and then by selecting households within these groupings with adults aged between 18 and 59 years. This sampling approach facilitates the extrapolation of results at the Metropolitan Lima level and the country’s urban and rural areas. For a deeper understanding of VIANEV’s methodology, referring to the survey’s technical report and previous studies is encouraged ^(13)^.

Entries missing essential variables for determining CVR were omitted from the analysis. The study was limited to participants aged 40 to 59 years to ensure the validity of comparisons across different scoring scales.

### Variables and Measurement

#### The primary variable was the calculation of CVR using four calculators

- AHA/ASCVD (American Heart Association/American College of Cardiology Atherosclerotic Cardiovascular Disease) ^(6)^: Designed for adults aged 40 to 79, this calculator evaluates the 10-year risk of atherosclerotic cardiovascular events. It includes age, gender, systolic blood pressure, hypertension treatment, total cholesterol, HDL cholesterol, type 2 diabetes mellitus (T2DM), and smoking habits.
- WHO Laboratory-Based Risk Chart ^(7)^: This World Health Organization calculator is based on risk factors requiring laboratory data. It applies to men and women aged 40 to 79 and considers variables like age, sex, smoking, systolic blood pressure, total cholesterol, and T2DM to predict the 10-year risk of myocardial infarction or stroke.
- WHO Non-Laboratory-Based Risk Chart ^(7)^: Also developed by the World Health Organization, this version does not require laboratory data and is suitable for the same age group as the laboratory-based version. It uses variables such as age, sex, smoking, and systolic blood pressure to estimate cardiovascular risk.
- Framingham Cardiovascular Risk (CVR Framingham) ^(8)^: The Framingham calculator estimates the risk of cardiovascular events over ten years for individuals aged 30 to 74. It considers factors like age, sex, systolic blood pressure, hypertension treatment, total cholesterol, HDL cholesterol, diabetes, and smoking.

The risk percentage of each study individual was calculated, categorizing the CVR from each calculator into low (< 10%) vs moderate/high (≥ 10%).

Potential associated factors considered were sex (male, female), age group (categorized into 40 to 49 years, and 50 to 59 years), educational level (up to primary, secondary, high), marital status (single, with a partner), natural region (Coast, mountains, and jungle), area of residence (urban, rural), alcohol consumption in the last year (yes, no), smoking status (never smoked, former smoker, current smoker), socioeconomic status (poor, not poor), presence of HTN (systolic blood pressure ≥ 140 and/or diastolic blood pressure ≥ 90, or if HTN is self-reported), presence of type 2 diabetes mellitus (T2DM) (fasting glucose ≥ 126 mg/dl or if T2DM is self-reported), physical activity (low, medium, high), nutritional status measured by body mass index (BMI = weight (kg)/height (meters)2) in average weight (BMI < 24.99 kg/m2), overweight (BMI ≥ 25 kg/m2 to BMI < 30 kg/m2), and obesity (≥30 kg/m2), abdominal obesity measured by waist circumference (WC) (WC ≥ 102 in males or ≥ 88 in females), and altitude (categorized from 0 to 1499 and 1500 to more).

### Data Collection and Procedure

Blood pressure was measured using a digital sphygmomanometer. Initially, the patient’s dominant arm was selected for the measurement. Then, two blood pressure readings were taken, and both averages were calculated for a preliminary diagnosis. In cases where a difference of 20 mmHg in systolic blood pressure (SBP) readings or ten mmHg in diastolic blood pressure (DBP) readings between the first and second measurement was observed, a third measurement was performed. Only measurements without such discrepancies were recorded. Blood pressure was predominantly taken in the morning, between 6:00 and 9:00 a.m.; if not possible, it was done between 7:00 and 9:00 p.m.

Patients were required to fast for 9 to 12 hours before laboratory sample collection. For these analyses, serum was drawn and transported under a cold chain to determine the lipid profile. Triglyceride levels were measured by an automatic end-point enzymatic-colorimetric method, while blood glucose levels were determined using previously calibrated portable glucometers (HemoCue Glucose 201 RT). An automated direct enzymatic colorimetric method determined HDL and total cholesterol levels.

Body mass index was calculated by dividing weight in kilograms by the square of height in meters. Waist circumference was measured at the end of expiration with a tape measure while the patient stood upright, bare torso and feet spaced between 25 and 30 cm at the upper edge of the iliac crest. This measure was taken three consecutive times, and the average of these measurements was considered the result.

### Statistical Analysis

Initially, a descriptive analysis of the sample will be conducted, where frequencies and percentages for categorical variables will be calculated, and measures of central tendency and dispersion (mean and standard deviation) for continuous variables will be obtained. This step will allow a basic understanding of the distribution of the interest variables in the studied population. Subsequently, a Poisson regression model with robust variance will be applied to evaluate the factors related to the concordance between the CVR calculators.

Finally, a concordance analysis, including Cohen’s kappa coefficient calculation, will be conducted to determine the degree of agreement between the different CVR calculators. This analysis will be complemented by a Venn Diagram visualizing the overlap between the calculators regarding risk classification.

### Ethical Aspects

This article was prepared using datasets from the VIANEV Survey, publicly available online. All personal identifiers were removed from these datasets before their publication, thus ensuring the anonymity of the patients.

Since this work was based on analyzing anonymous pre-existing information collected by third parties, submitting it for ethics committee evaluation was deemed unnecessary. This procedure aligns with ethical guidelines for health research, which specify that analyses of publicly accessible datasets without identifiable details do not require formal ethical review. However, precautionary measures were established to ensure that the study was conducted ethically and respectfully, preserving the rights and dignity of the participants in the original research.

## Results

The level of moderate/high CVD risk according to the type of calculator used was 5.44%, 4.73%, 6.62%, and 23.88% for WHO laboratory-based, WHO non-laboratory-based, AHA/ASCVD, and Framingham, respectively. Females comprised 57.21% of the sample, 68.09% lived on the coast, and 65.48% were in urban areas. Regarding harmful habits, 71.63% consumed alcohol in the last year, and 8.98% were current smokers. The prevalence of obesity was 33.33% and 54.76%, according to BMI and WC, respectively. The entire sample description is seen in Table 1.

**Table 1.**
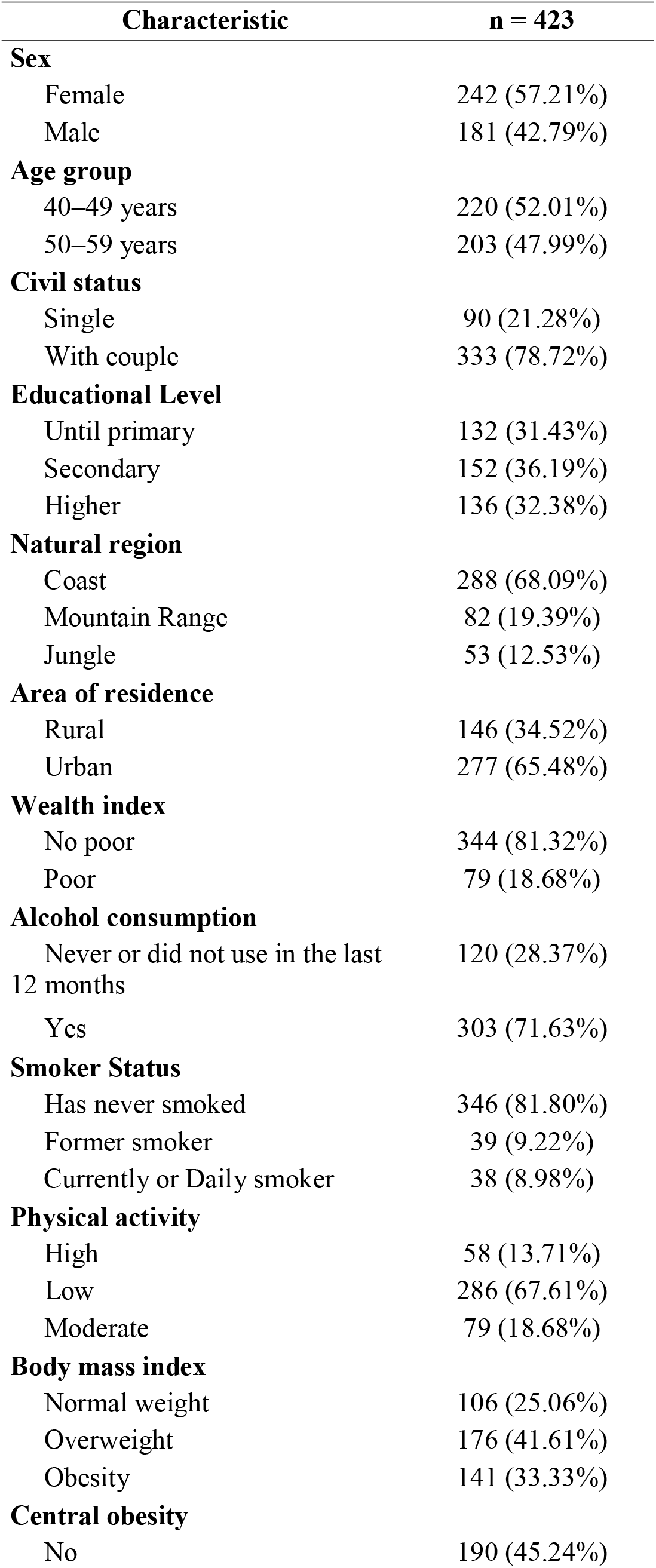
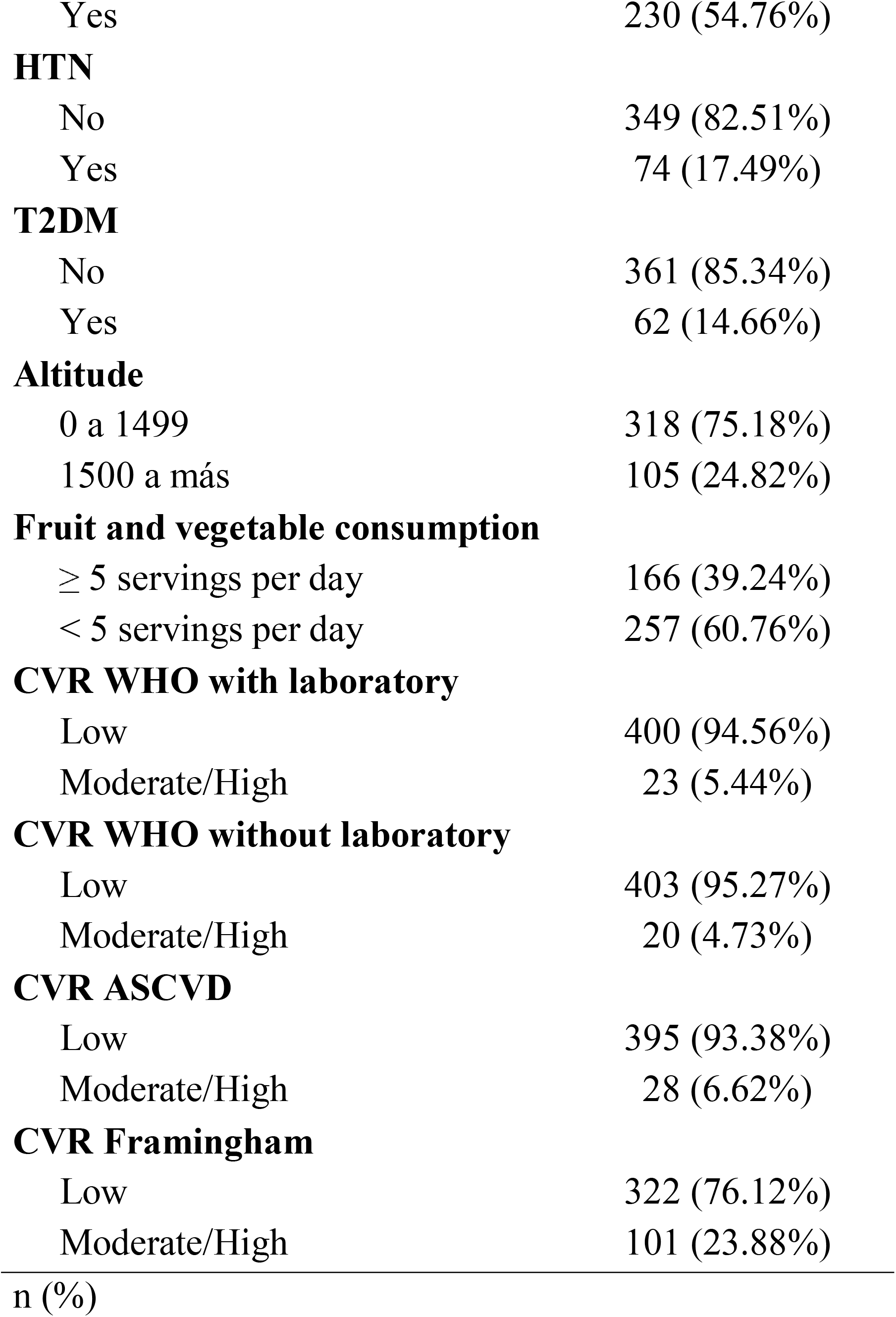
Demographic and anthropometric characteristics of participants.

**Table 2.**
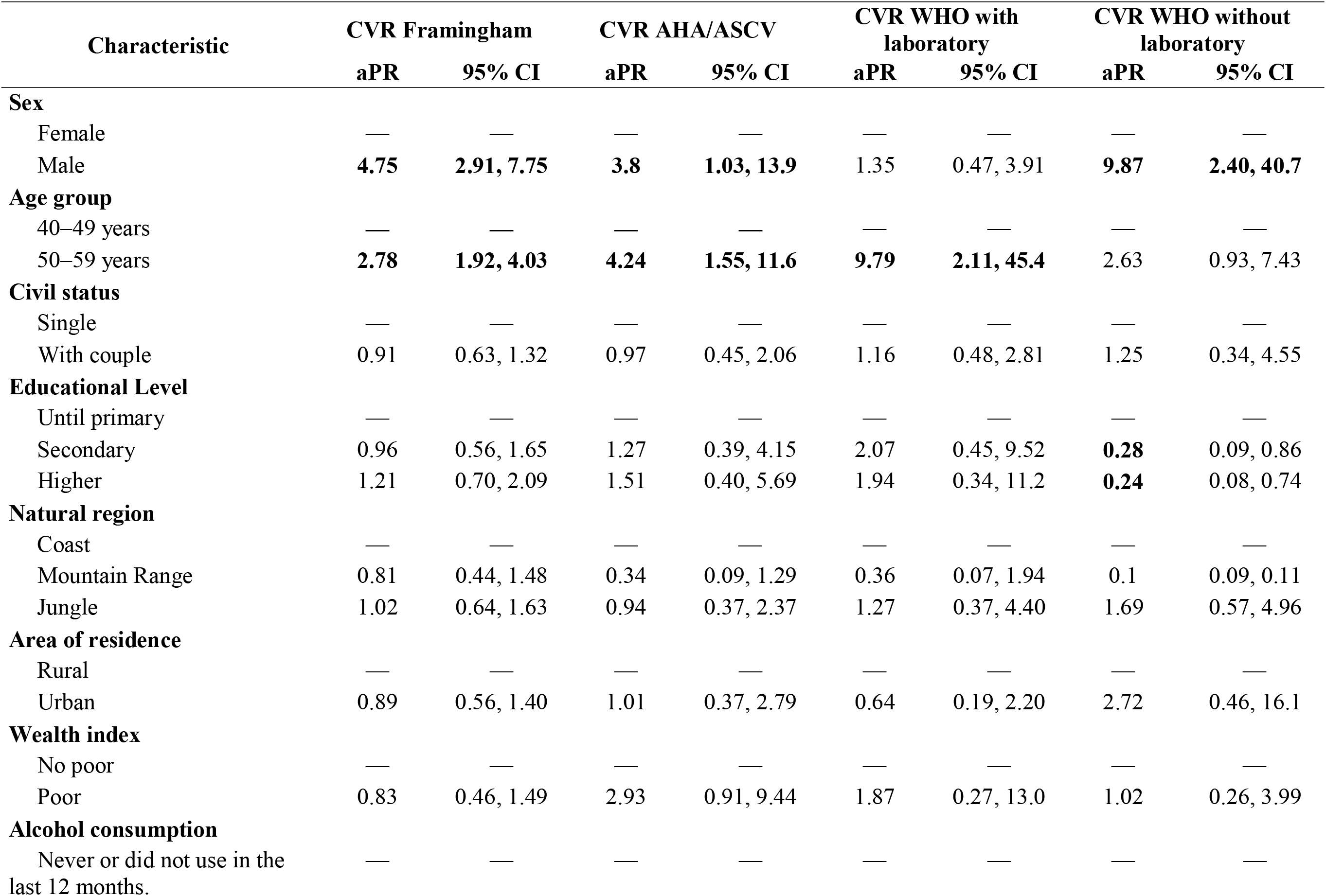
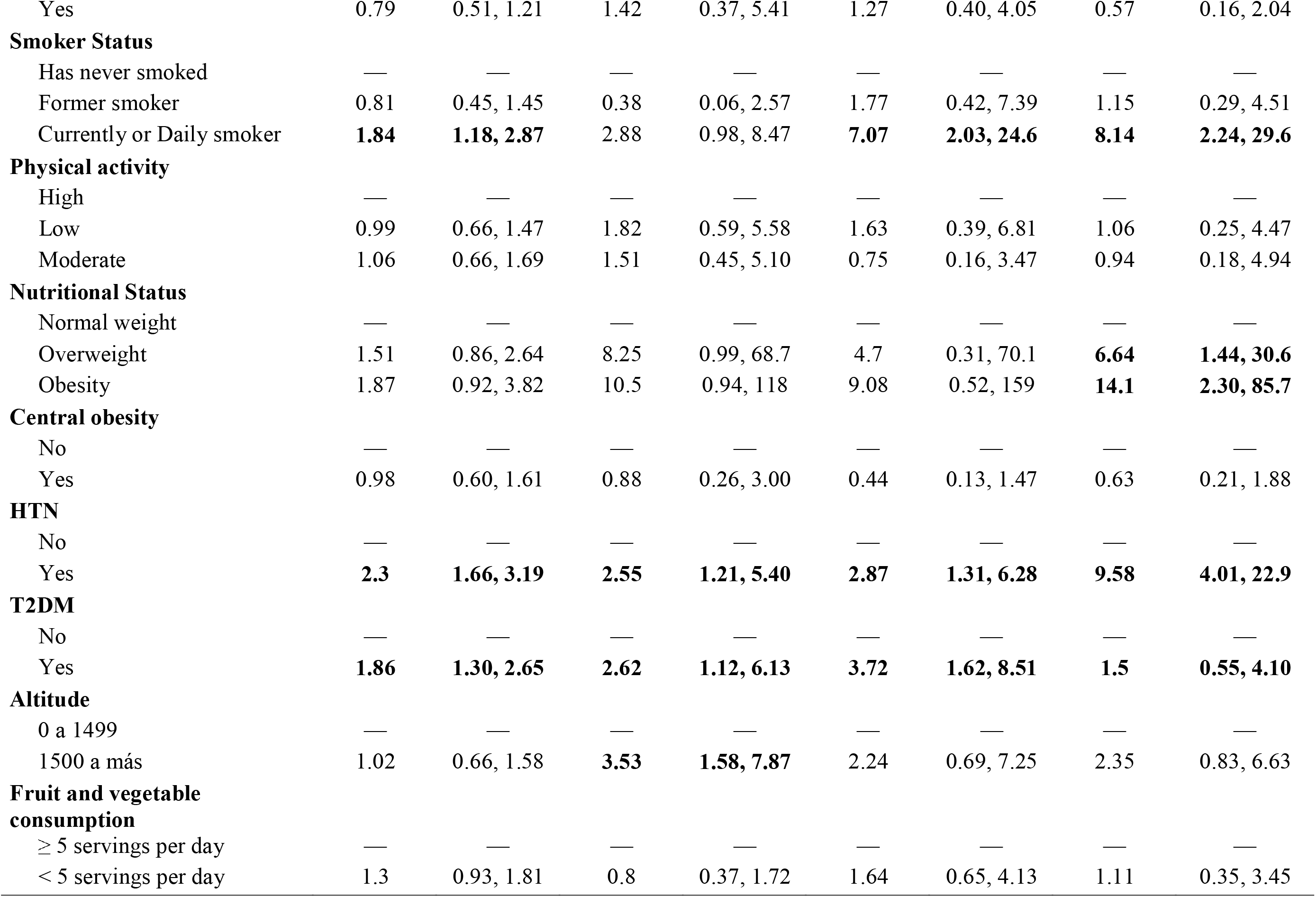
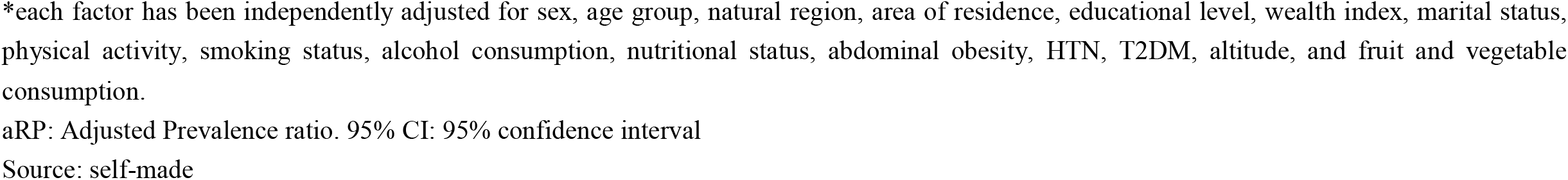
Multivariable regression analysis of the factors associated with CVR according to the calculators: WHO laboratory-based, WHO non-laboratory-based, AHA/ASCVD y, Framingham.

| Characteristic | CVR Framingham |  | CVR AHA/ASCVD |  | CVR WHO with laboratory |  | CVR WHO without laboratory |  |
| --- | --- | --- | --- | --- | --- | --- | --- | --- |
|  | aPR | 95% CI | aPR | 95% CI | aPR | 95% CI | aPR | 95% CI |
| **Sex** |  |  |  |  |  |  |  |  |
| Female | — | — | — | — | — | — | — | — |
| Male | **4.75** | **2.91, 7.75** | **3.8** | **1.03, 13.9** | 1.35 | 0.47, 3.91 | **9.87** | **2.40, 40.7** |
| **Age group** |  |  |  |  |  |  |  |  |
| 40–49 years | — | — | — | — | — | — | — | — |
| 50–59 years | **2.78** | **1.92, 4.03** | **4.24** | **1.55, 11.6** | **9.79** | **2.11, 45.4** | 2.63 | 0.93, 7.43 |
| **Civil status** |  |  |  |  |  |  |  |  |
| Single | — | — | — | — | — | — | — | — |
| With couple | 0.91 | 0.63, 1.32 | 0.97 | 0.45, 2.06 | 1.16 | 0.48, 2.81 | 1.25 | 0.34, 4.55 |
| **Educational Level** |  |  |  |  |  |  |  |  |
| Until primary | — | — | — | — | — | — | — | — |
| Secondary | 0.96 | 0.56, 1.65 | 1.27 | 0.39, 4.15 | 2.07 | 0.45, 9.52 | **0.28** | 0.09, 0.86 |
| Higher | 1.21 | 0.70, 2.09 | 1.51 | 0.40, 5.69 | 1.94 | 0.34, 11.2 | **0.24** | 0.08, 0.74 |
| **Natural region** |  |  |  |  |  |  |  |  |
| Coast | — | — | — | — | — | — | — | — |
| Mountain Range | 0.81 | 0.44, 1.48 | 0.34 | 0.09, 1.29 | 0.36 | 0.07, 1.94 | 0.1 | 0.09, 0.11 |
| Jungle | 1.02 | 0.64, 1.63 | 0.94 | 0.37, 2.37 | 1.27 | 0.37, 4.40 | 1.69 | 0.57, 4.96 |
| **Area of residence** |  |  |  |  |  |  |  |  |
| Rural | — | — | — | — | — | — | — | — |
| Urban | 0.89 | 0.56, 1.40 | 1.01 | 0.37, 2.79 | 0.64 | 0.19, 2.20 | 2.72 | 0.46, 16.1 |
| **Wealth index** |  |  |  |  |  |  |  |  |
| No poor | — | — | — | — | — | — | — | — |
| Poor | 0.83 | 0.46, 1.49 | 2.93 | 0.91, 9.44 | 1.87 | 0.27, 13.0 | 1.02 | 0.26, 3.99 |
| **Alcohol consumption** |  |  |  |  |  |  |  |  |
| Never or did not use in the last 12 months. | — | — | — | — | — | — | — | — |
| Yes | 0.79 | 0.51, 1.21 | 1.42 | 0.37, 5.41 | 1.27 | 0.40, 4.05 | 0.57 | 0.16, 2.04 |
| **Smoker Status** |  |  |  |  |  |  |  |  |
| Has never smoked | — | — | — | — | — | — | — | — |
| Former smoker | 0.81 | 0.45, 1.45 | 0.38 | 0.06, 2.57 | 1.77 | 0.42, 7.39 | 1.15 | 0.29, 4.51 |
| Currently or Daily smoker | **1.84** | **1.18, 2.87** | 2.88 | 0.98, 8.47 | **7.07** | **2.03, 24.6** | **8.14** | **2.24, 29.6** |
| **Physical activity** |  |  |  |  |  |  |  |  |
| High | — | — | — | — | — | — | — | — |
| Low | 0.99 | 0.66, 1.47 | 1.82 | 0.59, 5.58 | 1.63 | 0.39, 6.81 | 1.06 | 0.25, 4.47 |
| Moderate | 1.06 | 0.66, 1.69 | 1.51 | 0.45, 5.10 | 0.75 | 0.16, 3.47 | 0.94 | 0.18, 4.94 |
| **Nutritional Status** |  |  |  |  |  |  |  |  |
| Normal weight | — | — | — | — | — | — | — | — |
| Overweight | 1.51 | 0.86, 2.64 | 8.25 | 0.99, 68.7 | 4.7 | 0.31, 70.1 | **6.64** | **1.44, 30.6** |
| Obesity | 1.87 | 0.92, 3.82 | 10.5 | 0.94, 118 | 9.08 | 0.52, 159 | **14.1** | **2.30, 85.7** |
| **Central obesity** |  |  |  |  |  |  |  |  |
| No | — | — | — | — | — | — | — | — |
| Yes | 0.98 | 0.60, 1.61 | 0.88 | 0.26, 3.00 | 0.44 | 0.13, 1.47 | 0.63 | 0.21, 1.88 |
| **HTN** |  |  |  |  |  |  |  |  |
| No | — | — | — | — | — | — | — | — |
| Yes | **2.3** | **1.66, 3.19** | **2.55** | **1.21, 5.40** | **2.87** | **1.31, 6.28** | **9.58** | **4.01, 22.9** |
| **T2DM** |  |  |  |  |  |  |  |  |
| No | — | — | — | — | — | — | — | — |
| Yes | **1.86** | **1.30, 2.65** | **2.62** | **1.12, 6.13** | **3.72** | **1.62, 8.51** | **1.5** | **0.55, 4.10** |
| **Altitude** |  |  |  |  |  |  |  |  |
| 0 a 1499 | — | — | — | — | — | — | — | — |
| 1500 a más | 1.02 | 0.66, 1.58 | **3.53** | **1.58, 7.87** | 2.24 | 0.69, 7.25 | 2.35 | 0.83, 6.63 |
| **Fruit and vegetable consumption** |  |  |  |  |  |  |  |  |
| ≥ 5 servings per day | — | — | — | — | — | — | — | — |
| < 5 servings per day | 1.3 | 0.93, 1.81 | 0.8 | 0.37, 1.72 | 1.64 | 0.65, 4.13 | 1.11 | 0.35, 3.45 |

\*each factor has been independently adjusted for sex, age group, natural region, area of residence, educational level, wealth index, marital status, physical activity, smoking status, alcohol consumption, nutritional status, abdominal obesity, HTN, T2DM, altitude, and fruit and vegetable consumption.
aRP: Adjusted Prevalence ratio, 95% CI: 95% confidence interval
Source: self-made

According to the adjusted multivariable analysis in Table 3, factors commonly associated across almost all calculators were male sex, age between 50 – 59 years, current smoker, HTN, and T2DM. Isolated associations were found with educational level (WHO non-laboratory-based), obesity according to BMI (WHO non-laboratory-based), and altitude higher than 1500 meters above sea level (AHA/ASCVD).

**Table 3:** Concordance of CVR according to the calculators: WHO laboratory-based, WHO non-laboratory-based, AHA/ASCVD y Framingham.

| Test | Kappa | Agreement | Expected agreement | p-value |
| --- | --- | --- | --- | --- |
| WHO laboratory-based & WHO non-laboratory-based | 0.3876 | 94.09% | 90.35% | < 0.001 |
| WHO laboratory-based & AHA/ASCVD | 0.7289 | 96.93% | 88.66% | < 0.001 |
| WHO laboratory-based & Framingham | 0.3098 | 81.56% | 73.28% | < 0.001 |
| WHO non-laboratory-based & AHA/ASCVD | 0.3826 | 93.38% | 89.28% | < 0.001 |
| WHO non-laboratory-based & Framingham | 0.2732 | 80.85% | 73.65% | < 0.001 |
| AHA/ASCVD & Framingham | 0.3687 | 82.74% | 72.66% | < 0.001 |

The Venn diagram in Figure 1 illustrates the concordance among the CVD risk classifications of four different tools. Each oval represents one of the calculators, and the intersections between them show the number of patients classified in the same risk category by multiple calculators. All four calculators classify 77.4% of the observations in the same risk category. On the other hand, the WHO non-laboratory-based calculator uniquely identifies 12 patients (2.9% of the total evaluated), meaning these patients were only classified as at risk by this tool and not by the other three. In contrast, Framingham alone does not identify individuals with the unique risk that the rest of the calculators cannot detect.

**Figure 1.**
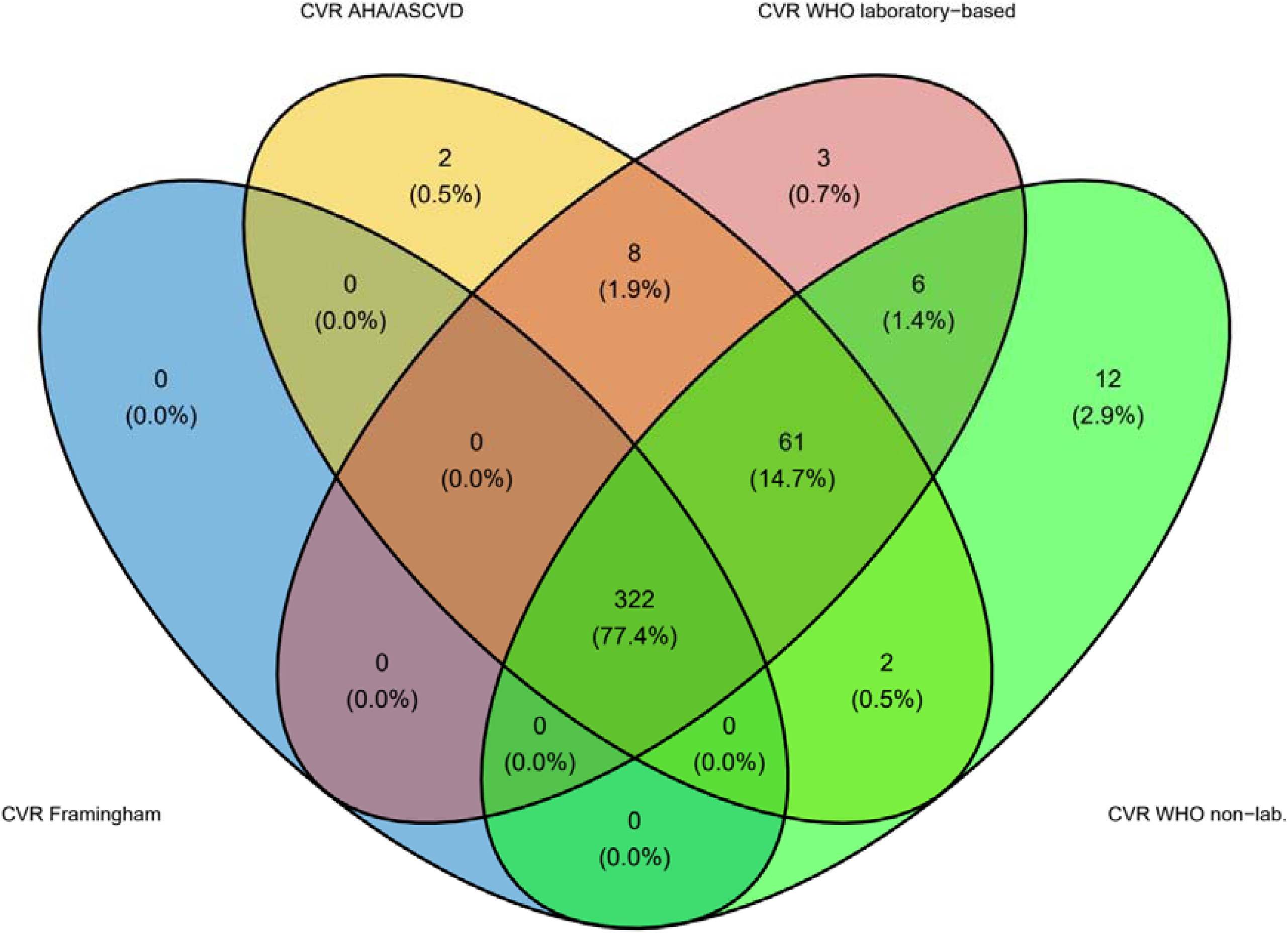
Venn diagram of RCV calculators.

Regarding the concordance analysis, all pairs of calculators present p-values < 0.001, indicating they are statistically significant. However, the strength of concordance varies, with the WHO laboratory-based and AHA/ASCVD combination showing the highest concordance (kappa of 0.7289). In contrast, concordance between WHO laboratory-based and Framingham and between WHO non-laboratory-based and Framingham is relatively lower (kappa of 0.3098 and 0.2732, respectively).

## Discussion

The marked difference in the prevalence and concordance of moderate/high CVD risk detected by the Framingham calculator compared to the WHO and AHA/ASCVD calculators could be due to various factors inherent to each tool’s design and specific algorithms.

The Framingham calculator, which showed a moderate/high-risk prevalence of 23.88%, is based on a set of risk variables that differs from the rest by including biochemical values such as total cholesterol and HDL cholesterol. These factors might be weighted differently in the Framingham algorithm than the other calculators, resulting in a higher sensitivity to identifying at-risk individuals ^(11,12,15–17)^. If we are looking at a population with a higher prevalence of dyslipidemias, this would increase the probability of classifying a person as high risk, especially in populations with specific demographic and health profiles like those found in Peru ^(18)^. Additionally, the initial population on which these prediction models were based might influence their applicability to other populations. The Framingham model was developed using data from a predominantly Caucasian population in the United States and may not perfectly adjust to populations with different epidemiological profiles, such as the Peruvian population, which may have different risk factor prevalences and patterns of cardiovascular diseases ^(8,19)^.

On the other hand, the WHO and AHA/ASCVD cardiovascular risk calculators are based on methodologies that reflect a global concern for preventing cardiovascular diseases ^(6,7)^. The WHO calculators were developed to be internationally applicable, with specific versions adapted to different geographic regions, considering the availability or absence of laboratory data ^(7)^. These tools use standard variables such as age, sex, blood pressure, and smoking habit but also adapt to reflect disease profiles and prevalent risk factors in different population contexts ^(20)^. The AHA/ASCVD has been recommended for use above other calculators ^(21,22)^.

The higher concordance observed between the WHO laboratory-based and AHA/ASCVD calculators could be attributed to the inclusion and similar weighting of typical clinical and biochemical risk factors, such as cholesterol levels and diabetes, which are key indicators of cardiovascular health. Both calculators require laboratory data to assess these factors, which may lead to a more accurate risk estimation in populations where such data are available and reliable. Moreover, both tools have been adjusted and validated using large databases that include a wide range of risk profiles, which may contribute to their consistency when applied to diverse populations ^(9)^.

The WHO non-laboratory-based calculator, designed for use in settings where laboratory tests are unavailable, means that while it can be applied more broadly, its ability to identify risks is based on a more limited spectrum of clinical variables. This could explain why it uniquely identifies a small proportion of people that the other calculators do not detect. The absence of biomarkers in its algorithm can lead to differences in risk classification, especially in individuals whose only significant risk factors are biochemical ^(23–25)^.

Interestingly, despite the Framingham calculator identifying more individuals at moderate/high risk, it does not uniquely identify any individuals compared to others. This may reflect a limitation in the Framingham calculator’s universal applicability, highlighting the need to adapt risk prediction tools to the specific characteristics of each population ^(26,27)^.

The analysis of risk factors associated with CVD risk revealed notable consistency among the different calculators in identifying male sex, older age (specifically between 50 and 59 years), smoking, HTN, and T2DM as significant predictors of increased CVD risk. This consistency underscores the universal relevance of these factors in cardiovascular risk stratification, reflecting the robustness of the calculators in capturing the critical determinants of CVD risk in different populations. The influence of these factors has been consistently shown in the literature to contribute to the development and progression of cardiovascular diseases, regardless of the geographical or demographic context ^(28–30)^.

However, our study also identified specific associations of factors with certain calculators. For example, educational level and obesity, as measured through BMI, were significantly associated with the risk calculated by the WHO non-laboratory-based tool. A variety of studies support the association between social determinants, lifestyle factors, and cardiovascular risk, particularly in the context of non-laboratory-based risk assessment tools. Khandia et al. ^(17)^ and Borhanuddin et al. ^(31)^ highlight the importance of factors such as age, weight, and cholesterol in predicting cardiovascular risk. Martínez-García et al. ^(32)^ and Stevens ^(7)^ et al. further emphasize the role of social and environmental factors in this context. Czeczelewski et al. ^(33)^ and Demirbaş and Kutlu ^(34)^ provide evidence of the relationship between body composition and cardiometabolic risk factors, with the latter suggesting that BMI alone may not be sufficient to predict these risks. Finally, the systematic review by Barbaresko et al. ^(35)^ underscores the importance of healthy lifestyle habits and the identification of risk factors, respectively, to reduce cardiovascular risk.

Moreover, the association of risk with altitude higher than 1500 meters above sea level, observed only in the AHA/ASCVD calculator, suggests that specific environmental conditions and possibly physiological adaptations to altitude may influence cardiovascular risk estimation. This finding is particularly relevant for countries with mountainous regions, like Peru, where the population may experience unique effects of altitude on cardiovascular health ^(36)^.

These differences in factor associations with the calculators highlight the importance of considering individual and contextual characteristics in cardiovascular risk assessment. It also underscores the need to adapt CVD risk prediction tools to reflect variations more accurately in risk profiles associated with specific sociodemographic, behavioral, and environmental factors.

### Contribution to the Field

This study highlights the pressing need to ensure cardiovascular risk calculators are validated and tailored to their application among distinct groups if required. Comparing the concordance and differences in risk classification among several internationally recognized calculators provides a solid foundation for understanding how these tools can be optimized to better reflect the risk profiles of populations with unique characteristics, like the Peruvian population. This adaptation is crucial for improving the accuracy of cardiovascular risk estimates and effectively preventing cardiovascular diseases in diverse geographical and cultural contexts.

By determining which cardiovascular risk factors disproportionately affect those living in Peru, including issues such as obesity, tobacco usage, and high-altitude conditions, public health officials can gain meaningful understandings that help plan initiatives to improve community wellness. By highlighting these specific factors, the study underscores the need for locally adapted prevention and management strategies to address the determinants of cardiovascular health in Peru more effectively. This personalized approach is crucial for implementing public health policies that can mitigate the impact of these conditions on the global burden of cardiovascular diseases.

The study contributes to the field of public health by fostering greater awareness of the importance of cardiovascular prevention. Demonstrating variations in risk estimates among different calculators and highlighting the influence of modifiable risk factors reinforces the message that early prevention and lifestyle modification are potent tools against cardiovascular diseases. This awareness can drive public health campaigns to educate the population about cardiovascular risks and promote healthy lifestyle habits.

Finally, the study offers valuable insights for improving strategies for assessing and managing cardiovascular risk. It provides evidence that can guide the selection of risk assessment tools in clinical practice. This is particularly relevant for health professionals in Peru and similar regions. They can benefit from a deeper understanding of choosing and using these calculators to inform clinical decisions and public health policies to prevent cardiovascular diseases in their communities.

### Study Limitations

While the cross-sectional design limits the capacity to ascertain causality between identified danger elements and cardiovascular hazard, this research is meaningfully confined by its transversal nature, which prohibits concluding whether risk determinants cause elevated risk or vice versa. While the analyses offer valuable insights into how diverse factors interrelate with CVD risk among Peruvians, they alone cannot decisively conclude that these risk factors directly induce greater chances of CVD without uncertainty. Another significant limitation is the potential lack of adaptation of the cardiovascular risk calculators used for the specific study population. While this study endeavors to juxtapose and assess the suitability of these instruments in Peru, it remains plausible that the calculators, formulated in divergent demographic and health settings, may fail to completely encapsulate the intricacies of the cardiovascular risk portrayal of Peru’s inhabitants. This could influence the accuracy of risk estimates and, therefore, the intervention recommendations based on these tools.

While self-reported details concerning certain risk factors like smoking practices or chronic illness history could allow for misrepresentations to infiltrate the data, relying solely on such information may potentially skew the results. Additionally, while measures were taken to ensure consistency and plausibility of blood pressure measurements, secondary data limits the ability to verify the accuracy of all health measurements included in the analysis. Data quality variability could undermine the dependability of the links uncovered between dangerous elements and cardiovascular disease hazards if not addressed. Ultimately, though this research offers meaningful revelations regarding heart disease danger among Peruvians, simply extending these results to other groups demands prudent restraint. Differences in epidemiological profiles, prevalent risk factors, and socioeconomic and environmental conditions may limit the direct applicability of the results to contexts outside Peru. It would be prudent to conduct similar studies in diverse populations to validate and possibly adjust cardiovascular risk estimation tools according to the specific needs of each population.

## Conclusions

There are discrepancies in CVD risk levels according to the different calculators evaluated, especially with the Framingham calculator, underscoring the need for a more personalized approach to risk estimation. Although common risk factors such as male gender, age range of 50 to 59 years, smoking, arterial hypertension, and diabetes were identified, these variations suggest that prediction tools might not be universally applicable in their current form.

Longitudinal studies are recommended to address these limitations and improve accuracy in predicting CVD risk. These studies would confirm causal relationships between identified risk factors and CVD risk and better understand how cardiovascular risk evolves in response to changes in risk factors. Such research would provide robust evidence for adapting or designing new risk calculators calibrated explicitly for the Peruvian population.

Furthermore, it’s essential to consider calibrating existing risk calculators or developing new tools that more accurately reflect the epidemiological and health profiles of the Peruvian population. This would involve adjusting calculators based on local data and considering the genetic, environmental, and social particularities influencing cardiovascular risk in Peru. Implementing these recommendations would significantly improve risk stratification and, thereby, the prevention and management of cardiovascular diseases in this population.

## Data Availability

The data supporting the findings of this study can be accessed by the original research paper at the follow link: https://www.datosabiertos.gob.pe/dataset/estado-nutricional-en-adultos-de-18-59-a%C3%B1os-per%C3%BA-2017-%E2%80%93-2018.

## Acknowledgments

A special thanks to the members of the Tropical Diseases Research Institute, Universidad Nacional Toribio Rodríguez de Mendoza de Amazonas (UNTRM), Amazonas, Peru, for their support and contributions throughout the completion of this research.

## Informed consent

It was not necessary to obtain informed consent in this Study

## Author contributions

**Víctor Juan Vera-Ponce:** Methodology, Data Analysis, Writing – Review & Editing.

**Fiorella E. Zuzunaga-Montoya:** Conceptualization, Data Analysis, Writing – Review & Editing.

**Joan A. Loayza-Castro:** Data Analysis, Methodology, Validation, Writing – Original Draft.

**Luisa Erika Milagros Vásquez Romero:** Data Analysis, Methodology, Validation, Writing – Original Draft.

**Carmen Inés Gutierrez De Carrillo:** Validation, Visualization, Project Administration, Supervision, Methodology, Writing – Review & Editing.

**Enrique Vigil-Ventura:** Supervision, Methodology, Funding Acquisition, Writing – Review & Editing.

